# Copy number heterogeneity identifies ER+ breast cancer patients with adverse outcome after adjuvant endocrine therapy

**DOI:** 10.1101/2021.11.15.21266346

**Authors:** Tom van den Bosch, Oscar M Rueda, Carlos Caldas, Louis Vermeulen, Daniël M Miedema

**Author notes:** Correspondence: Daniël M. Miedema, Meibergdreef 9, 1105 AZ Amsterdam, The Netherlands.

## Abstract

**Background:** Endocrine therapy forms the backbone of adjuvant treatment for estrogen-receptor positive (ER+) breast cancer. However, it remains unclear whether adjuvant treatment improves survival rates in low-risk patients. Low intra-tumor heterogeneity (ITH) has been shown to confer low-risk for recurrent disease. Here, it is studied if chromosomal copy number ITH (CNH) can identify low-risk ER+, lymph node-negative breast cancers patients who do not benefit from adjuvant endocrine therapy.

**Methods:** Lymph node-negative ER+ patients from the METABRIC dataset were retrospectively analyzed (n=708). CNH was determined from a single bulk copy number measurement for each patient. Patients were stratified by CNH score as low, medium or high. Overall survival (OS) was compared between patients that did, or did not receive adjuvant endocrine therapy per CNH group with Cox proportional-hazards models, using propensity score weights to correct for confounders.

**Results:** Adjuvant endocrine therapy improved the relapse free survival (RFS) for high-CNH/high-risk patients treatment (Hazard Ratio [HR] = 0.55, 95% Confidence Interval [CI] = 0.35 to 0.87), but not for low CNH/risk patients treatment (HR = 0.88, 95% CI = 0.50 to 1.55). For low-CNH/low-risk patients adjuvant endocrine therapy was associated with impaired OS (HR = 1.62, 95% CI = 1.10 to 2.40). Multivariable Cox analysis identified a significant interaction between CNH and endocrine therapy for OS (HR = 0.77, 95% CI = 0.62 to 0.96).

**Conclusions:** This retrospective study of lymph node-negative, ER+ breast cancer finds that patients identified as low-risk using CNH display reduced OS when treated with adjuvant endocrine therapy.

---

Breast cancer is the most common malignancy among women worldwide. The majority of breast cancers are estrogen receptor positive (ER+) and detected at an early stage, for which endocrine therapy is the cornerstone of adjuvant treatment [1]. Adjuvant endocrine therapies are highly effective at reducing recurrences of ER+ breast cancers. Tamoxifen reduces the recurrence rates with about 50% during the first ten years after start of treatment and significantly reduces mortality [2]; Aromatase inhibitors, which were introduced later, further reduce recurrence rates by 30% compared to treatment with Tamoxifen alone [3].

Novel strategies to further improve survival of ER+ breast cancer patients typically rely on intensifying adjuvant therapy. Through extending the duration of endocrine treatment from 5 to 10 years [4-6], by administering chemotherapeutics to patients identified as high-risk [7-9] or by identifying and tackling resistance mechanisms [10]. Yet, the majority of patients who did not receive any adjuvant endocrine treatment are free from recurrence 10 years after diagnosis [2]. In particular, patients with lymph node-negative disease have a reasonable prognosis, with 65% of patients free from recurrence after ten years, despite not receiving adjuvant endocrine treatment.

Upfront identification of low-risk patients is of key clinical importance, as long-term treatment with endocrine therapy comes with severe side-effects. Endocrine therapy is associated with hot-flushes, sexual dysfunction, weight gain and fatigue, which all severely reduce the quality-of-life [11]. Potentially life-threatening, although not frequently observed, side-effects of endocrine therapy include endometrial cancer and pulmonary embolism [6, 11]. For patients with low-risk of recurrence the side-effects of extended treatment with endocrine therapy hence might outweigh the benefit from the therapy [12]. Indeed, de-escalation is considered in treatment guidelines of early-stage breast cancers [13]. Furthermore, individualized risk models that guide treatment decisions are widely used to estimate the benefit of adjuvant endocrine therapy, but so far only incorporate limited molecular features (e.g. Predict) [14].

Intra-tumor heterogeneity (ITH) is a pan-cancer marker for survival and strongly associates with risk of recurrence [15]. Recent studies demonstrated extensive ITH at the copy-number level in breast cancers, which associated with survival [16-18]. To assess the impact of chromosomal copy number ITH (CNH) in large scale studies, we recently introduced a tool to infer CNH from a single sample copy number profile [19]. In the present study, we used copy number and clinical data from the METABRIC cohort [20, 21] to study if CNH can stratify risk in ER+ node-negative breast cancer patients and whether low-risk patients identified by CNH benefit from adjuvant endocrine therapy.

## Methods

### Study population and data source

We selected ER+ and lymph node-negative breast cancer patients from the METABRIC cohort [20] (Figure 1). Patients treated with (neo)adjuvant chemotherapy were excluded from analyses, such that included patients received either endocrine therapy or no systemic therapy as adjuvant treatment. Clinical data of the METABRIC cohort was obtained from the supplementary information of Rueda e*t al*. [21]. Patients for which clinical data was incomplete were excluded from the analysis. Copy number variation data were obtained from cBioPortal. Patients for which no copy number data was available were excluded from the analyses.

**Figure 1.**
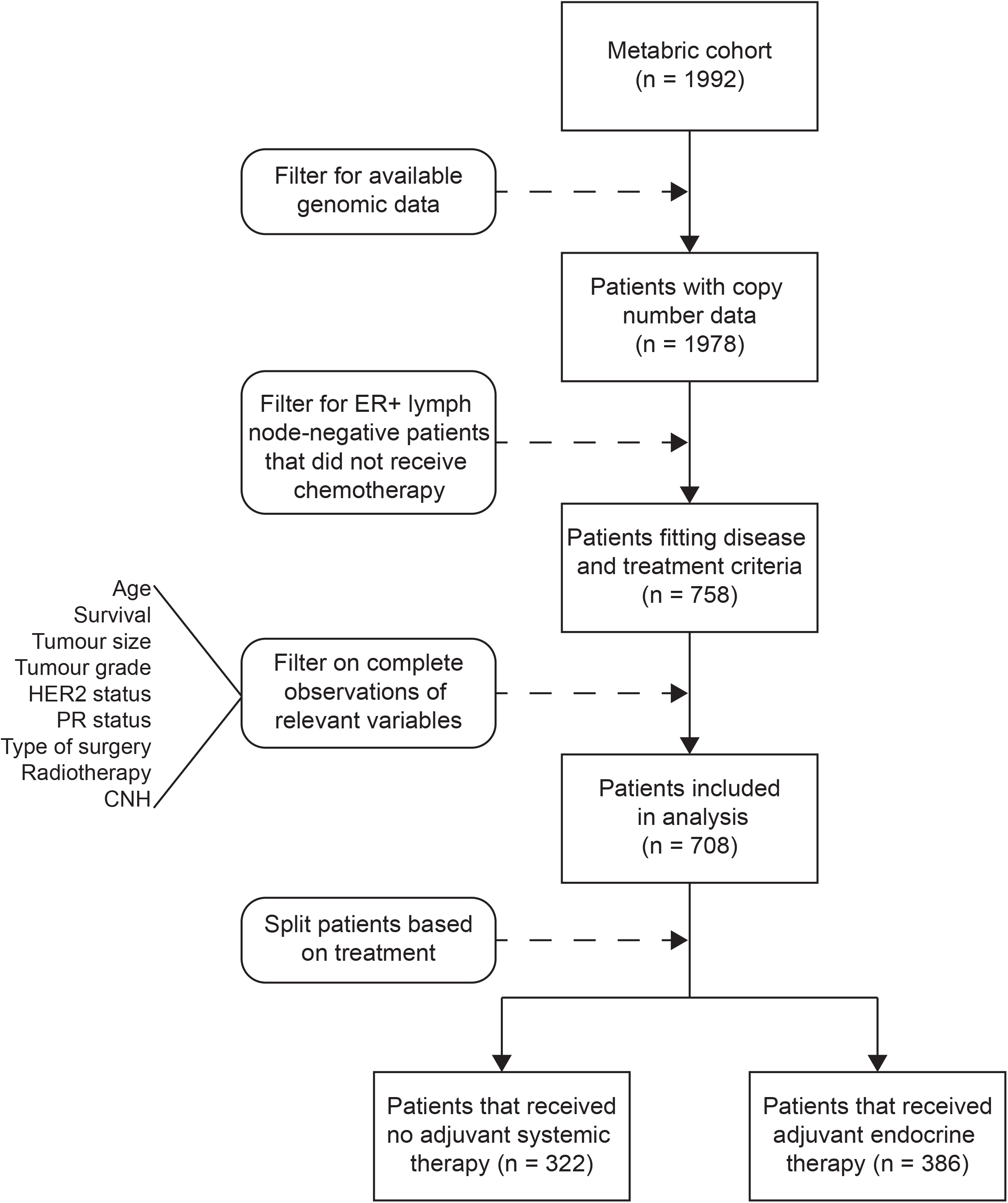
Flowchart of patient selection for the present study.

### CNH inference

CNH was inferred from a single bulk measurement of copy number variants as described previously [19]. In short, a grid search of ploidies (1.5, 1.55,.., 5) and purities (0.2, 0.21,.., 1) was performed to transform relative segmented copy numbers to absolute copy numbers. In the grid search, the distance of absolute copy numbers to the closest integer values was measured for each segment, and the weighted average over all segments was determined. CNH was identified as the minimum of the weighted distances of absolute copy numbers to integer values.

### Statistical analyses

All patients were sorted into 3 CNH groups (CNH/risk low, medium and high) of equal size based on the rank-ordered CNH. Patient groups were defined by CNH group and treatment regimen (adjuvant endocrine therapy or not). Differences in overall survival (OS), disease specific survival (DSS) and recurrence free survival (RFS) were analyzed between groups.

The Kaplan-Meier method was used to visualize survival curves of the CNH groups and the log-rank test was applied to test for statistical differences, for patients treated with endocrine therapy or not separately.

Baseline characteristics of patients receiving adjuvant endocrine therapy or not were compared using Fisher’s exact test for binary variables and a chi-square test for non-binary categorical variables. Continuous variables were translated to categorical variables (Age: <60, 60-69, 70-79 and 80+; Tumor size: <=20mm, >20mm).

To compare outcomes after adjuvant endocrine treatment or not for patients of similar CNH, the propensity score was determined as the probability of the received adjuvant endocrine treatment based on age, surgery type, PR status, HER2 status, tumor grade, tumor size and radiotherapy.

Univariate Cox proportional-hazards models were created for each CNH group to determine the hazard-ratios associated with endocrine treatment, using inverse probability of treatment weighting (IPW) [22, 23]. A weighted log-rank test was used to compare survival between groups treated with adjuvant endocrine therapy and untreated patients, for all three CNH groups.

Multivariable Cox proportional-hazard models including predictors: CNH as a continuous predictor, adjuvant endocrine treatment, surgery type, radiotherapy, age at diagnosis HER2 status, PR status, tumor grade, tumor size and the interaction between CNH and adjuvant endocrine therapy, were created to evaluate the prognostic value of CNH and the interaction between CNH and adjuvant endocrine therapy.

CNH was determined and Kaplan-Meier curves were generated using MATLAB R2021a. All other statistical analyses were done in R 3.6.1 using the packages: *survival, RISCA, weights* and *stats*.

## Results

Patients with ER+ and lymph node-negative disease that did not receive (neo)adjuvant chemotherapy and from which copy number data was available were selected from the METABRIC cohort (n = 708; Figure 1). Table 1 shows the baseline characteristics of patients included in the present study. The median age at diagnosis of patients in the present study was 63. All patients underwent surgery for breast cancer (mastectomy n = 367 or breast conserving n = 341), n = 367 patients were treated with radiotherapy and n = 386 received adjuvant endocrine therapy. Patients treated with adjuvant endocrine therapy were more likely to also receive radiotherapy treatment (P = 0.009, Fisher exact test), were on average older (P < 0.001, chi-square test) and had larger tumors (P < 0.001, fisher exact test) of higher grade (P < 0.001, chi-square test) than patients not treated with adjuvant endocrine therapy.

**Table 1.** Baseline characteristics of the patients in the present study.

|  |  | Patient received adjuvant endocrine therapy |  |  |
| --- | --- | --- | --- | --- |
|  | All | No | Yes | *P* |
| n | 708 (100) | 322 (45.5) | 386 (54.5) |  |
| Age groups |  |  |  | <0.001 |
| <60 | 289 (40.8) | 161 (50.0) | 128 (33.2) |  |
| 60-70 | 34 (4.8) | 17 (5.3) | 17 (4.4) |  |
| 70-80 | 217 (30.6) | 85 (26.4) | 132 (34.2) |  |
| 80+ | 168 (23.7) | 59 (18.3) | 109 (28.2) |  |
| Her2 status |  |  |  | 0.142 |
| negative | 666 (94.1) | 308 (95.7) | 358 (92.7) |  |
| positive | 42 (5.9) | 14 (4.3) | 28 (7.3) |  |
| PR status |  |  |  | 0.073 |
| negative | 196 (27.7) | 78 (24.2) | 118 (30.6) |  |
| positive | 512 (72.3) | 244 (75.8) | 268 (69.4) |  |
| Tumour size |  |  |  | <0.001 |
| <= 20mm | 292 (41.2) | 158 (49.1) | 134 (34.7) |  |
| >20mm | 416 (58.8) | 164 (50.9) | 252 (65.3) |  |
| Type of surgery |  |  |  | 0.195 |
| mastectomy | 367 (51.8) | 176 (54.7) | 191 (49.5) |  |
| breast conserving | 341 (48.2) | 146 (45.3) | 195 (50.5) |  |
| Tumour grade |  |  |  | <0.001 |
| 1 | 99 (14.0) | 50 (15.5) | 49 (12.7) |  |
| 2 | 377 (53.2) | 197 (61.2) | 180 (46.6) |  |
| 3 | 232 (32.8) | 75 (23.3) | 157 (40.7) |  |
| Patient received radiotherapy |  |  |  | 0.009 |
| no | 341 (49.2) | 173 (53.7) | 168 (43.5) |  |
| yes | 367 (51.8) | 149 (46.3) | 218 (56.5) |  |

For all patients CNH was inferred from a single copy number variation measurement as described previously [19]. Patients were rank ordered based on CNH, and three CNH groups of equal size were defined as: low CNH (CNH < 0.03), medium CNH (0.03 ≤ CNH < 0.076) and high CNH (0.076 ≤ CNH) (Figure 2A). For patients not treated with endocrine therapy CNH stratifies for survival, with low CNH corresponding to low risk of recurrence and high CNH corresponding to high risk of recurrence (Figure 2B and Supplementary Figure 1A,B). For patients treated with adjuvant endocrine therapy no relation between CNH and survival was observed (Figure 2C and Supplementary Figure 1C,D). These results suggested that CNH is prognostic for survival, but more importantly also indicated an interaction between CNH and efficacy of adjuvant endocrine therapy.

**Figure 2.**
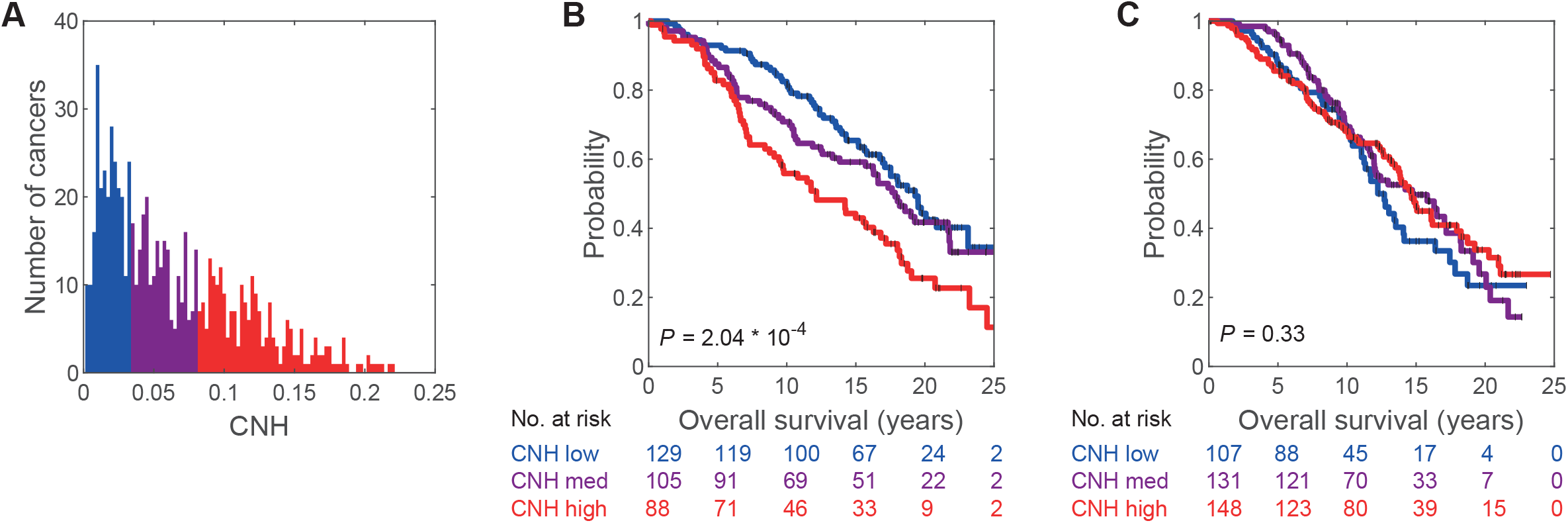
Distribution of copy number heterogeneity (CNH) and overall survival (OS) per CNH group and treatment. A) Distribution of CNH with definition of CNH groups: blue, CNH-low; purple, CNH-medium; red, CNH-high. B) Kaplan-Meier curves for patients that received no adjuvant systemic therapy per CNH group. C) Kaplan-Meier curves for patients that received adjuvant endocrine therapy per CNH group. Reported P-values are from comparison of CNH-low and CNH-high groups.

As this cohort is not randomized for adjuvant endocrine therapy, direct comparison of outcomes between treatment strategies for patients of similar CNH requires balancing of baseline characteristics between treatment groups. To this end, the inverse probability of treatment was calculated according to the propensity score [22, 23]. Weighting by the propensity score resulted in balanced patient groups treated and untreated by endocrine therapy, with no significant differences between baseline characteristics (Supplementary Table 1 and Supplementary Figure 2).

The results from IPW weighted univariate Cox proportional-hazards models and the weighted log-rank method applied to the propensity weighted data are shown in Figure 3 and Supplementary Figure 3. For patients with high CNH the relapse free survival (RFS; HR = 0.55, 95 % CI = 0.35 to 0.87) and disease specific survival (DSS; HR = 0.61, 95 % CI = 0.38 to 0.97) were both significantly improved by adjuvant endocrine therapy. A trend towards better OS for patients with high CNH treated with adjuvant endocrine therapy was observed (HR = 0.71, 95% CI = 0.51 to 1.00). In contrast, for patients with low CNH adjuvant endocrine therapy neither improved RFS (HR = 0.88, 95% CI = 0.55 to 1.55) nor DSS (HR = 1.31, 95% CI = 0.67 to 2.57). Strikingly, the OS for patients with low CNH was worse when treated with adjuvant endocrine therapy compared to no systemic adjuvant treatment (HR = 1.62. 95% CI = 1.10 to 2.40). The observed HR’s of adjuvant endocrine treatment for patients with medium CNH were between those of patients with high CNH and low CNH for all outcomes.

**Figure 3.**
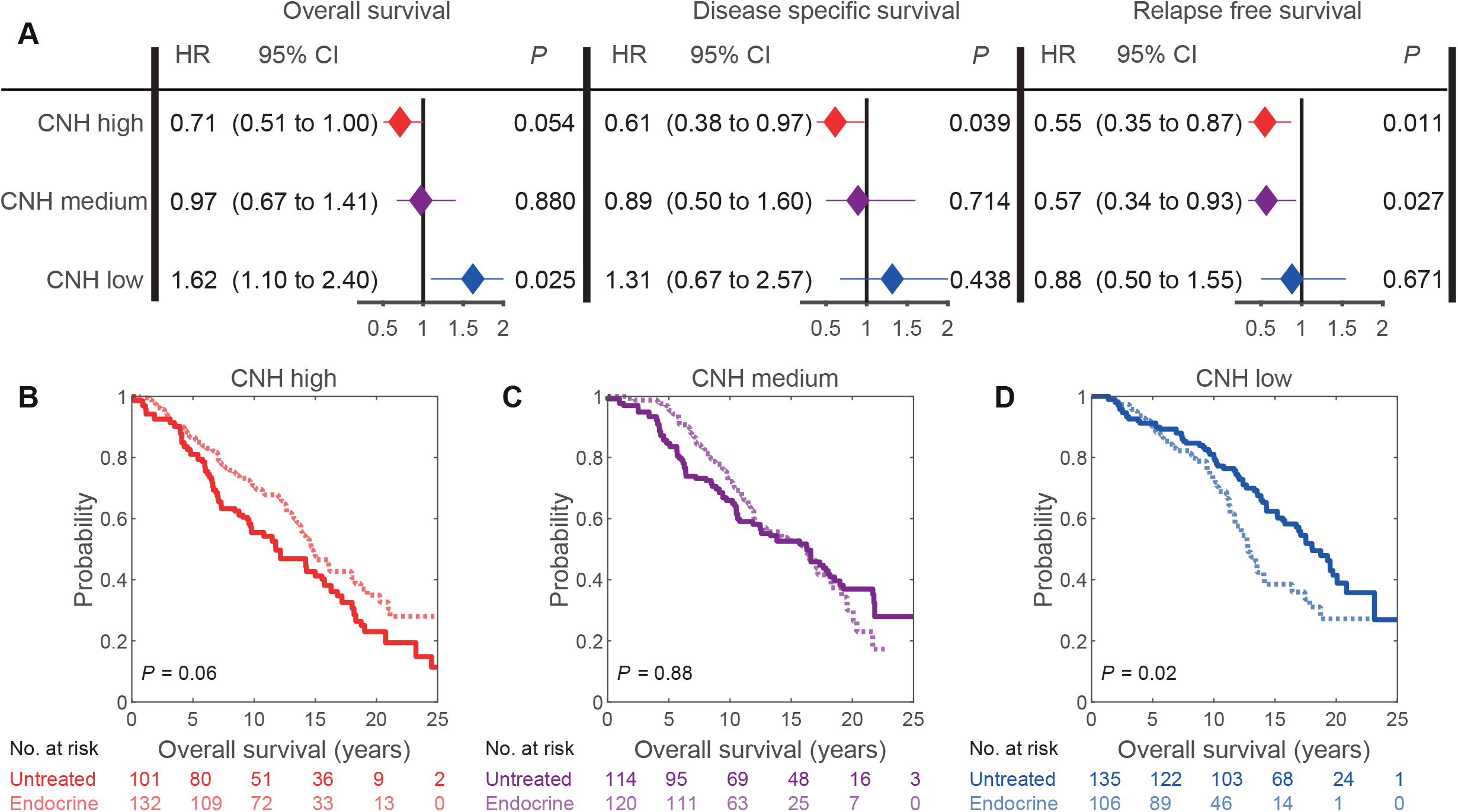
Forest plots and Kaplan-Meier curves of propensity score weighted patient groups. A) Hazard ratio’s for adjuvant endocrine therapy vs. no adjuvant systemic therapy for different suvival outcomes in a univariate setting. B-D) Propensity score weighted Kaplan-Meier curves for OS in CNH-high (B), CNH-medium (C) and CNH-low (D) patient groups.

A multivariable Cox proportional-hazards model was constructed to formally address the interaction between CNH and adjuvant endocrine therapy. To arrive at a single interaction variable between CNH and adjuvant endocrine therapy, CNH was taken as continuous variable normalized by the mean and standard deviation. The model outcomes for are shown in Table 2. Adjuvant endocrine therapy significantly improved RFS (HR = 0.64, 95% CI = 0.47 to 0.86). High CNH was associated with poor survival for all outcomes (RFS: HR = 1.30, 95% CI = 1.08 to 1.56 DSS: HR = 1.55, 95% CI = 1.26 to 1.90; OS: HR = 1.28, 95% CI = 1.10 to 1.49). The interaction between CNH and endocrine therapy was negative for all outcomes, but only significant for OS (HR = 0.77, 95% CI = 0.62 to 0.96).

**Table 2.**
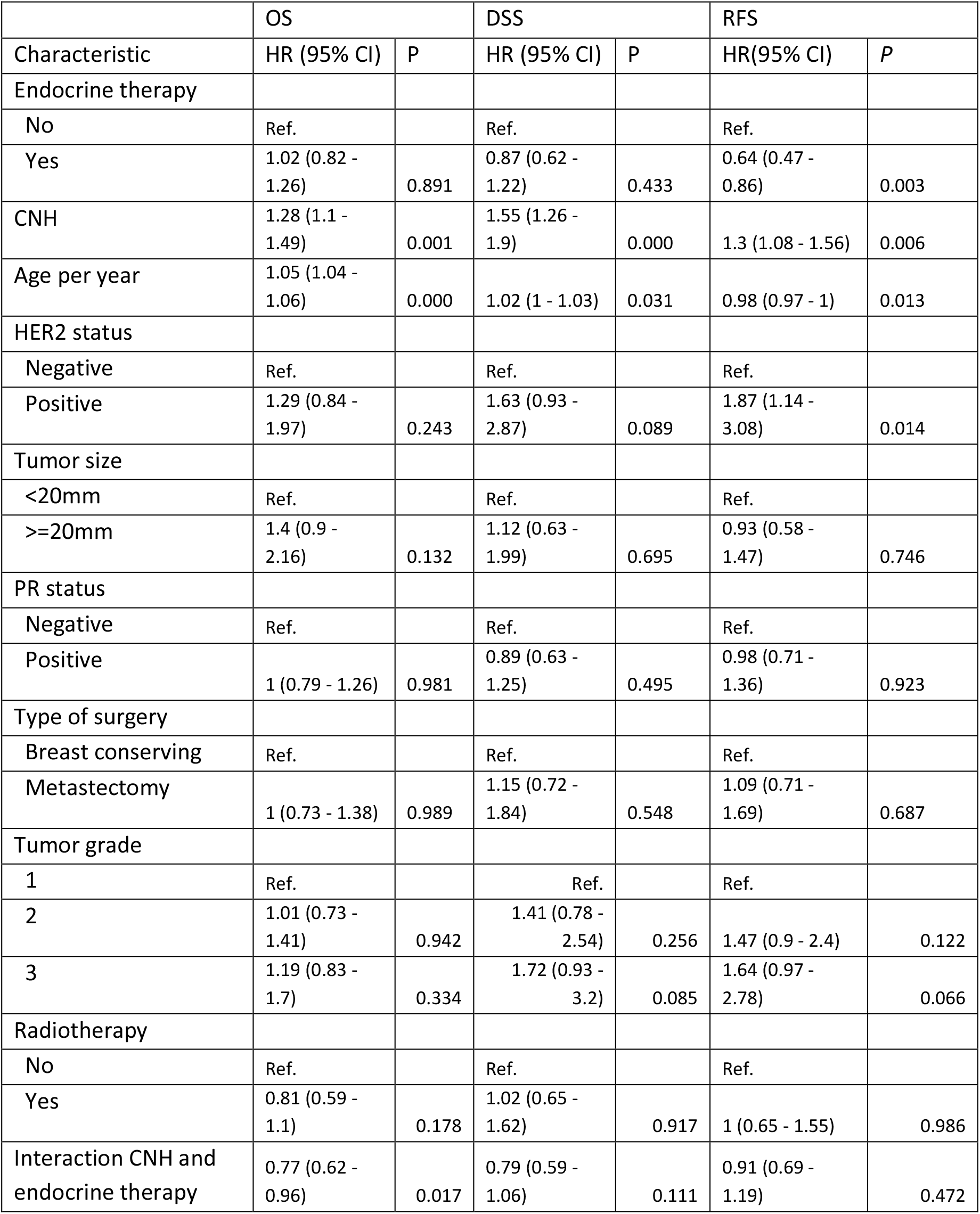
Multivariable Cox proportional-hazards for overall, disease specific and relapse free survival.

## Discussion

In this retrospective analysis of ER+, lymph-node negative breast cancer patients it was observed that the ∼33% patients with the lowest genetic risk have worse OS when receiving adjuvant endocrine therapy compared to no endocrine treatment. Genetic risk was assessed by the level of ITH in chromosomal copy numbers (i.e. CNH) and was inferred from a single measurement per patient [19]. The results presented suggest that patients with high genetic risk defined by CNH benefit from adjuvant endocrine treatment, while for patients with low genetic risk adjuvant endocrine therapy might be omitted.

Adverse outcome after a treatment seems counter-intuitive at first glance. In cancers from virtually all types there is a trend toward combining more therapeutics, which is thought to improve treatment efficacy by synergy or simple additive effects [24]. Yet, systemic treatments often come with severe side-effects and widespread overtreatment with endocrine therapy was recently suggested [25]. Moreover, many breast cancer patients do not die directly from their cancers [26]. For patients with low baseline risk the harm inflicted by side-effects, such as pulmonary embolism and endometrial cancer associated with years long treatment with adjuvant endocrine therapy, hence might outweigh the benefit from the treatment.

Various approaches to identify low genetic risk patients in early-stage ER+ breast cancers for treatment selection have been proposed previously. A range of commercial and academic tools have been developed to identify low-risk ER+ breast cancer patients who might not need (neo)adjuvant chemotherapy on top of endocrine therapy [7-9, 27-29]; or to dissect patients who benefit from extended treatment with adjuvant endocrine therapy (10 years) from low risk patients for whom 5 year treatment with endocrine therapy is sufficient [5]. Recently, a tool was introduced to identify low-risk patients who might not need adjuvant endocrine therapy from a cohort of untreated patients [30]. From patients that were not treated with adjuvant systematic therapy a subgroup with excellent prognosis could be identified. However, as that study did not include patients treated with endocrine therapy, it is not clear if the outcome of these low-risk patients could be further improved by adjuvant endocrine therapy. The novelty of the current study is that it identified a significant group of ER+ patients with low genetic-risk that have an impaired OS associated with adjuvant endocrine therapy, in comparison to patients with similar genetic risk that received no adjuvant systemic therapy.

Genetic risk was identified in the present study from the level of ITH. The level of ITH is a well-established pan-cancer indicator of genetic risk with a clear biological rationale: a cancer with high genetic diversity is more likely to contain escaping clones. Previous tools used to identify low genetic risk in early stage breast cancer are obtained by identifying the biological tumor characteristics that differ between good and poor responders or survivors. Although effective, these tools miss an *a priori* rationale that can explain why low risk patients are at low risk [31]. A further difference between previous studies and the current approach is that the latter is based on copy number variations, while the former are based on gene and/or protein expression levels.

The present study has several limitations. First, the current work is a retrospective analysis of data from the observational METABRIC cohort. Differences in baseline patient, tumor and treatment characteristics between treatment groups were corrected for using propensity to treatment scores. However, treatment with adjuvant endocrine therapy increased over the years and differed in frequency between the sub-cohorts of METABRIC. These factors could not be corrected for using propensity to treatment scores without disturbing the balancing of baseline patient and tumor characteristics. Second, the large majority of patients in the METABRIC cohort received only tamoxifen, while only a small proportion of patients received aromatase inhibitors. For post-menopausal women tamoxifen is switched to aromatase inhibitors after two to three years in present clinical practice. Aromatase inhibitors are therefore underrepresented in the current study. Third, it could be that some patients registered as not treated with endocrine therapy were actually treated with endocrine therapy but the treatment was not registered in the clinical data. This would suggest that the real interaction between CNH and endocrine therapy is possibly bigger than reported in the present study. Fourth, the benefit of adjuvant endocrine therapy observed in the METABRIC cohort is smaller than was reported from a meta-analysis of 20 trials of Tamoxifen for early breast cancer[2]. In the present cohort, we observed a significant benefit from endocrine therapy on RFS, but not for OS. The meta-analysis of the effect of Tamoxifen demonstrated a significant gain of both OS and RFS, although the effect for RFS was substantially larger than for OS [2]. Fifth, no validation on a second independent dataset has been performed yet.

Adjuvant endocrine therapy is currently prescribed for nearly all ER+ breast cancer patients. The results presented here suggest that in addition to expression of estrogen receptor, and clinical features such as included in Predict [14], the level of CNH can inform whether a breast cancer patient will benefit from adjuvant endocrine treatment. For up to 33% of ER+ lymph node-negative breast cancer patients adjuvant treatment with endocrine therapy might result in adverse OS. To verify these potentially clinically impactful results, further analysis of patients randomized for adjuvant treatment with endocrine therapy or not, is warranted.

## Data Availability

All clinical and copy number data is available in the publication of Rueda et al (Nature 2019, https://doi.org/10.1038/s41586-019-1007-8).
CNH values for patients in METABRIC and other data related to this study are available upon request.

https://www.ebi.ac.uk/ega/studies/EGAS00000000083

## Author contributions

TvdB, LV and DMM designed the study. TvdB, OMR, CC, LV and DMM analyzed data. TvdB, LV and DMM wrote the manuscript.

## Acknowledgements

We thank Dr. Erik van Dijk for useful discussions.

## Data availability

CNH values for patients in METABRIC and other data related to this study are available upon request.

## References

1. Burstein HJ. Systemic Therapy for Estrogen Receptor-Positive, HER2-Negative Breast Cancer. N Engl J Med 2020;383(26):2557–2570.

2. Early Breast Cancer Trialists’ Collaborative G, Davies C, Godwin J, et al. Relevance of breast cancer hormone receptors and other factors to the efficacy of adjuvant tamoxifen: patient-level meta-analysis of randomised trials. Lancet 2011;378(9793):771–84.

3. Early Breast Cancer Trialists’ Collaborative G. Aromatase inhibitors versus tamoxifen in early breast cancer: patient-level meta-analysis of the randomised trials. Lancet 2015;386(10001):1341–1352.

4. Burstein HJ, Temin S, Anderson H, et al. Adjuvant endocrine therapy for women with hormone receptor-positive breast cancer: american society of clinical oncology clinical practice guideline focused update. J Clin Oncol 2014;32(21):2255–69.

5. Davies C, Pan H, Godwin J, et al. Long-term effects of continuing adjuvant tamoxifen to 10 years versus stopping at 5 years after diagnosis of oestrogen receptor-positive breast cancer: ATLAS, a randomised trial. Lancet 2013;381(9869):805–16.

6. Goss PE, Ingle JN, Pritchard KI, et al. Extending Aromatase-Inhibitor Adjuvant Therapy to 10 Years. N Engl J Med 2016;375(3):209–19.

7. Cardoso F, van’t Veer LJ, Bogaerts J, et al. 70-Gene Signature as an Aid to Treatment Decisions in Early-Stage Breast Cancer. N Engl J Med 2016;375(8):717–29.

8. Gnant M, Sestak I, Filipits M, et al. Identifying clinically relevant prognostic subgroups of postmenopausal women with node-positive hormone receptor-positive early-stage breast cancer treated with endocrine therapy: a combined analysis of ABCSG-8 and ATAC using the PAM50 risk of recurrence score and intrinsic subtype. Ann Oncol 2015;26(8):1685–91.

9. Sparano JA, Gray RJ, Makower DF, et al. Adjuvant Chemotherapy Guided by a 21-Gene Expression Assay in Breast Cancer. New England Journal of Medicine 2018;379(2):111–121.

10. Hanker AB, Sudhan DR, Arteaga CL. Overcoming Endocrine Resistance in Breast Cancer. Cancer Cell 2020;37(4):496–513.

11. Franzoi MA, Agostinetto E, Perachino M, et al. Evidence-based approaches for the management of side-effects of adjuvant endocrine therapy in patients with breast cancer. Lancet Oncol 2021;22(7):e303–e313.

12. Pan H, Gray R, Braybrooke J, et al. 20-Year Risks of Breast-Cancer Recurrence after Stopping Endocrine Therapy at 5 Years. N Engl J Med 2017;377(19):1836–1846.

13. Curigliano G, Burstein HJ, Winer EP, et al. De-escalating and escalating treatments for early-stage breast cancer: the St. Gallen International Expert Consensus Conference on the Primary Therapy of Early Breast Cancer 2017. Ann Oncol 2017;28(8):1700–1712.

14. Wishart GC, Azzato EM, Greenberg DC, et al. PREDICT: a new UK prognostic model that predicts survival following surgery for invasive breast cancer. Breast Cancer Res 2010;12(1):R1.

15. McGranahan N, Swanton C. Clonal Heterogeneity and Tumor Evolution: Past, Present, and the Future. Cell 2017;168(4):613–628.

16. Baslan T, Kendall J, Volyanskyy K, et al. Novel insights into breast cancer copy number genetic heterogeneity revealed by single-cell genome sequencing. Elife 2020;9.

17. Minussi DC, Nicholson MD, Ye H, et al. Breast tumours maintain a reservoir of subclonal diversity during expansion. Nature 2021;592(7853):302–308.

18. Xu Z, Verma A, Naveed U, et al. Deep learning predicts chromosomal instability from histopathology images. iScience 2021;24(5):102394.

19. van Dijk E, van den Bosch T, Lenos KJ, et al. Chromosomal copy number heterogeneity predicts survival rates across cancers. Nat Commun 2021;12(1):3188.

20. Curtis C, Shah SP, Chin SF, et al. The genomic and transcriptomic architecture of 2,000 breast tumours reveals novel subgroups. Nature 2012;486(7403):346–52.

21. Rueda OM, Sammut SJ, Seoane JA, et al. Dynamics of breast-cancer relapse reveal late-recurring ER-positive genomic subgroups. Nature 2019;567(7748):399–404.

22. Austin PC. The performance of different propensity score methods for estimating marginal hazard ratios. Stat Med 2013;32(16):2837–49.

23. Austin PC, Stuart EA. Moving towards best practice when using inverse probability of treatment weighting (IPTW) using the propensity score to estimate causal treatment effects in observational studies. Stat Med 2015;34(28):3661–79.

24. Bayat Mokhtari R, Homayouni TS, Baluch N, et al. Combination therapy in combating cancer. Oncotarget 2017;8(23):38022–38043.

25. Derks MGM, Bastiaannet E, Kiderlen M, et al. Variation in treatment and survival of older patients with non-metastatic breast cancer in five European countries: a population-based cohort study from the EURECCA Breast Cancer Group. Br J Cancer 2018;119(1):121–129.

26. Chapman JA, Meng D, Shepherd L, et al. Competing causes of death from a randomized trial of extended adjuvant endocrine therapy for breast cancer. J Natl Cancer Inst 2008;100(4):252–60.

27. Buus R, Sestak I, Kronenwett R, et al. Comparison of EndoPredict and EPclin With Oncotype DX Recurrence Score for Prediction of Risk of Distant Recurrence After Endocrine Therapy. J Natl Cancer Inst 2016;108(11).

28. Sestak I, Buus R, Cuzick J, et al. Comparison of the Performance of 6 Prognostic Signatures for Estrogen Receptor-Positive Breast Cancer: A Secondary Analysis of a Randomized Clinical Trial. JAMA Oncol 2018;4(4):545–553.

29. Loi S, Haibe-Kains B, Desmedt C, et al. Predicting prognosis using molecular profiling in estrogen receptor-positive breast cancer treated with tamoxifen. BMC Genomics 2008;9:239.

30. Sjostrom M, Chang SL, Fishbane N, et al. Comprehensive Transcriptomic Profiling Identifies Breast Cancer Patients Who May Be Spared Adjuvant Systemic Therapy. Clin Cancer Res 2020;26(1):171–182.

31. Kern SE. Why your new cancer biomarker may never work: recurrent patterns and remarkable diversity in biomarker failures. Cancer Res 2012;72(23):6097–101.

