## Supplemental Table and Figures for "Copy number heterogeneity identifies ER+ breast cancer patients with adverse outcome after adjuvant endocrine therapy"

|  | Patient received adjuvant endocrine therapy | | |
| --- | --- | --- | --- |
|  | No | Yes | P |
| n | 350 (49.4) | 358 (50.6) |  |
| Age groups |  |  | 0.10664 |
| <60 | 147 (42.0) | 138 (35.8) |  |
| 60-70 | 23 (6.6) | 12 (3.1) |  |
| 70-80 | 96 (27.4) | 118 (30.6) |  |
| 80+ | 84 (24.0) | 90 923.3) |  |
| Her2 status |  |  | 0.68324 |
| negative | 331 (94.6) | 364 (94.3) |  |
| positive | 19 (5.4) | 22 (5.7) |  |
| PR status |  |  | 0.68917 |
| negative | 96 (27.4) | 130 (33.7) |  |
| positive | 254 (72.6) | 256 (66.3) |  |
| Tumour size |  |  | 0.68059 |
| <= 20mm | 148 (42.3) | 172 (44.6) |  |
| >20mm | 202 (57.7) | 214 (55.4) |  |
| Type of surgery |  |  | 0.79772 |
| mastectomy | 186 (53.1) | 185 (47.9) |  |
| breast conserving | 164 (46.9) | 201 (52.1) |  |
| Tumour grade |  |  | 0.77124 |
| 1 | 49 (14.0) | 49 (12.7) |  |
| 2 | 193 (55.1) | 190 (49.2) |  |
| 3 | 108 (30.9) | 120 (31.0) |  |
| Patient received radiotherapy |  |  | 0.74488 |
| no | 177 (50.6) | 201 (52.1) |  |
| yes | 173 (49.4) | 185 (47.9) |  |

**Supplementary Table 1**. Baseline characteristics after propensity score weighting of patients that received adjuvant endocrine therapy or not.

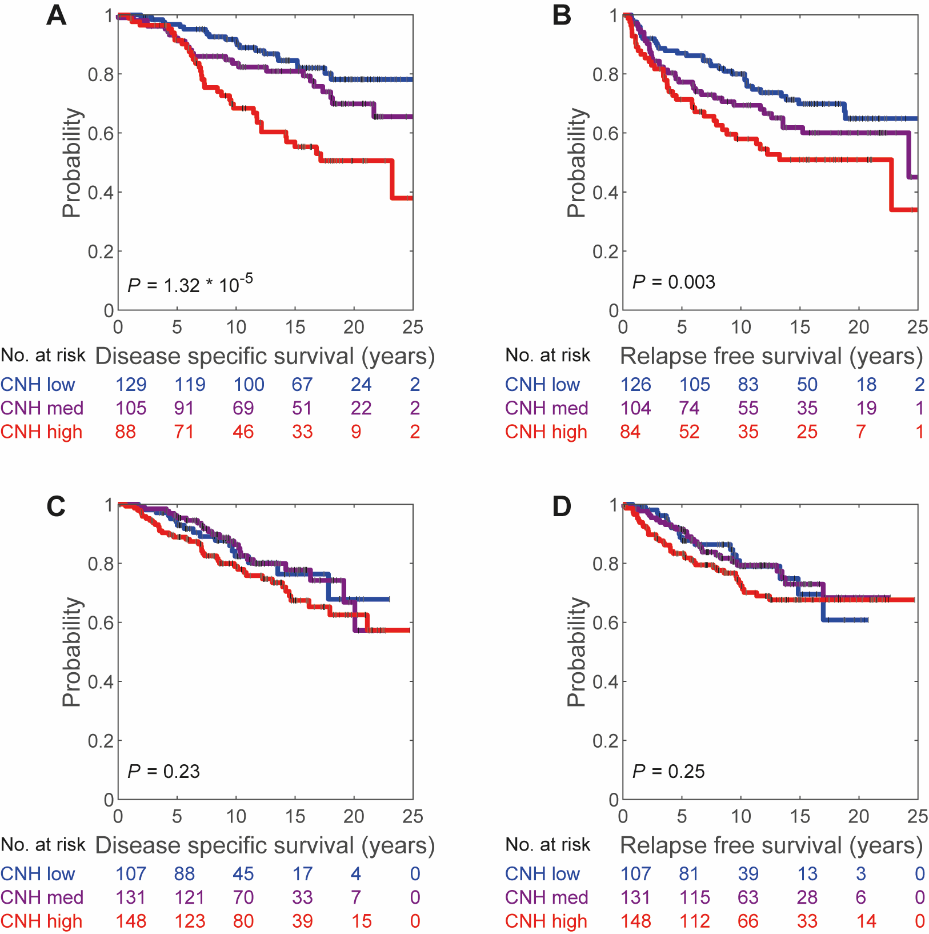

**Supplementary Figure 1**. Kaplan-Meier survival curves for disease specific and relapse free survival per treatment and CNH group. A) Disease specific survival of patients not treated with systemic adjuvant therapy. B) Relapse free survival of patients not treated with systemic adjuvant therapy. C) Disease specific survival of patients treated with adjuvant endocrine therapy. D) Relapse free survival of patients treated with adjuvant endocrine therapy. Reported P-values are from comparisons of CNH low and CNH high groups.

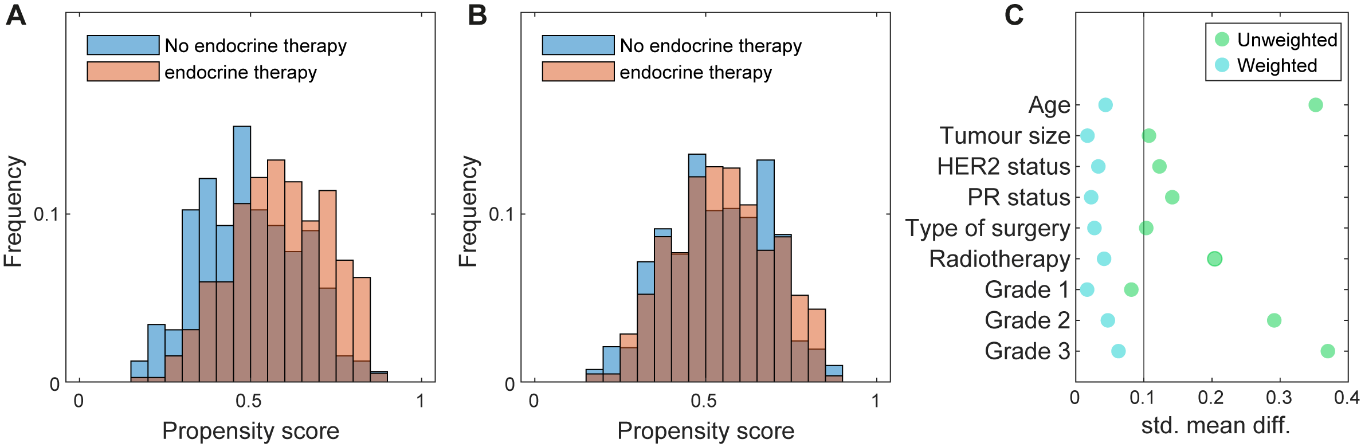

**Supplementary Figure 2**. Propensity scores of patients treated with adjuvant endocrine therapy or not. A) Distribution of propensity scores in unweighted population. B) Distribution of propensity scores after propensity score weighting. C) Standardized mean difference in baseline characteristics between adjuvant endocrine therapy treated or not patients, before and after propensity score weighting.

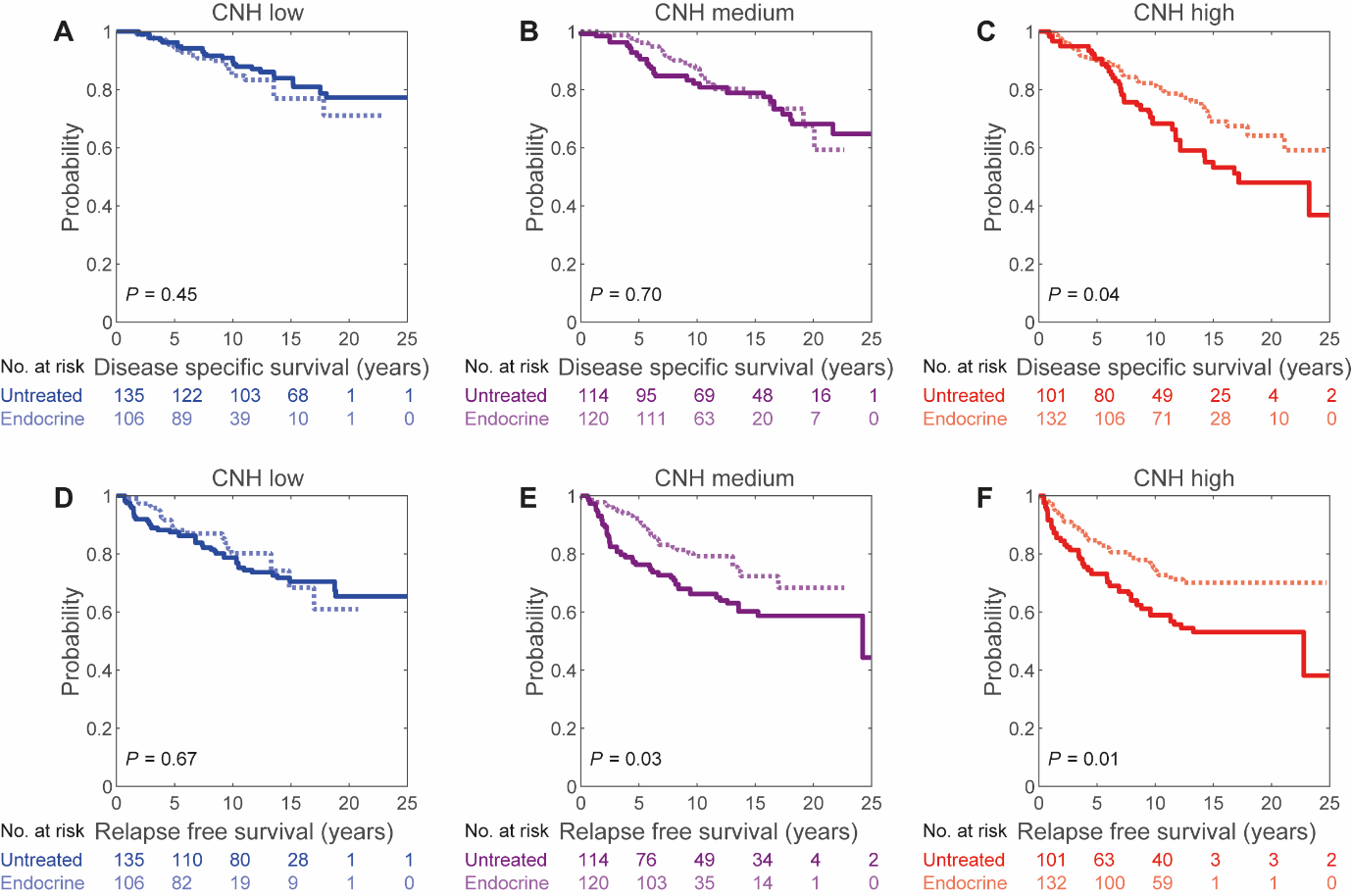

**Supplementary Figure 3**. Kaplan-Meier survival curves for propensity score weighted patient groups. A-C) Disease specific survival for patients with low CNH (A), medium CNH (B) and high CNH (C). D-F) Relapse free survival for patients with low CNH (D), medium CNH (E) and high CNH (F).
